# CLIF-Net: Intersection-guided Cross-view Fusion Network for Infection Detection from Cranial Ultrasound

**DOI:** 10.1101/2025.07.21.25331887

**Authors:** Mingzhao Yu, Mallory R. Peterson, Kathy Burgoine, Thaddeus Harbaugh, Peter Olupot-Olupot, Melissa Gladstone, Cornelia Hagmann, Frances M. Cowan, Andrew Weeks, Sarah U. Morton, Ronald Mulondo, Edith Mbabazi-Kabachelor, Steven J. Schiff, Vishal Monga

**Author notes:** M. Yu, M. Peterson and K. Burgoine contributed equally to this work. S. Schiff and V. Monga shared seniority.

## Abstract

This paper addresses the problem of detecting possible serious bacterial infection (pSBI) of infancy, i.e. a clinical presentation consistent with bacterial sepsis in newborn infants using cranial ultrasound (cUS) images. The captured image set for each patient enables multiview imagery: coronal and sagittal, with geometric overlap. To exploit this geometric relation, we develop a new learning framework, called the intersection-guided Cross-view Local-and Image-level Fusion Network (CLIF-Net). Our technique employs two distinct convolutional neural network branches to extract features from coronal and sagittal images with newly developed multi-level fusion blocks. Specifically, we leverage the spatial position of these images to locate the intersecting region. We then identify and enhance the semantic features from this region across multiple levels using cross-attention modules, facilitating the acquisition of mutually beneficial and more representative features from both views. The final enhanced features from the two views are then integrated and projected through the image-level fusion layer, outputting pSBI and non-pSBI class probabilities. We contend that our method of exploiting multi-view cUS images enables a first of its kind, robust 3D representation tailored for pSBI detection. When evaluated on a dataset of 302 cUS scans from Mbale Regional Referral Hospital in Uganda, CLIF-Net demonstrates substantially enhanced performance, surpassing the prevailing state-of-the-art infection detection techniques.

## I. INTRODUCTION

NEONATAL sepsis, traditionally defined as the occurrence of a systemic inflammatory response to infection resulting in symptoms within 28 days of birth, is typically identified through bacterial isolation from biological samples such as blood or cerebrospinal fluid (CSF) [1]. In the absence of recovery of causative pathogens, especially common in low-and middle-income countries (LMIC) settings, the clinical diagnosis is often made of presumed serious bacterial infection (pSBI). Globally, serious bacterial infections contribute to an estimated 21% of neonatal deaths and represent a primary cause of infant mortality in countries such as Uganda. Notably, neonatal pSBI may culminate in central nervous system (CNS) infection, provoking inflammation of the meninges (meningitis), ventricles (ventriculitis), or cerebrum (encephalitis).

Blood cultures from a neonate clinically diagnosed with pSBI and CSF cultures in neonates with suspected neurological involvement are the ‘gold standards’ for diagnosis [2]. However, in resource-limited settings, the availability of laboratory facilities is often scarce, making blood and CSF cultures infeasible options due to cost and time required to send samples to distant facilities. When the diagnosis of pSBI is uncertain, some neonates will be exposed to antibiotic therapy and other treatments that they do not need and have associated risks; other neonates will receive inadequate treatment, leading to increased risk of morbidity and mortality.

Cranial Ultrasound (cUS) is widely utilized as a low-cost, easily transportable, and simple-to-use imaging modality for identifying pre-and post-natal neurological abnormalities [3], [4]. It presents a plausible solution for the detection of CNS involvement in neonatal pSBI cases, thereby potentially improving detection and management without depending on CSF sampling and expensive laboratory facilities. The correlation between cUS findings and abnormal CSF analysis has been identified, demonstrating patterns indicative of CNS infection such as moderate to severe cortical and/or white matter (WM) echogenicities [5]. Other structural abnormalities include hyperechoic signals (without defined shape) that can represent inflammation and hypoechoic signals (without defined shape but often spherical in nature) that can represent cysts, etc. However, accurately identifying abnormalities in cUS findings necessitates neonatal medical experts, which presents an obstacle to widespread screening and point-of-care diagnosis. Thus, the development of an automated and accurate diagnosis system of pSBI with CNS involvement based on cUS images bears significant value. Traditional image classification methods often rely on manually designed feature extractors and classifiers [6], [7]. However, this approach can be challenging when applied to pSBI detection, because it necessitates in-depth knowledge from experienced experts. Moreover, the use of images in detecting pSBI is still an underexplored area. Deep learning techniques have demonstrated their power in extracting patterns from data. Such methods have consistently outperformed traditional computer vision techniques across a variety of image-related tasks [8]–[12].

Infection patterns in the brain are inherently threedimensional and may manifest differently across various imaging planes due to the distinct appearances of anatomical structures in each plane. Similarly, physicians employ multiple planes to corroborate the presence of pathologies such as infections. For instance, pSBI can be represented on cUS in patterns such as hyperechoic lining of the ventricles, which is referred to as ventriculitis. The ventricles can also contain inflammatory debris in the setting of infection. Both patterns are better appreciated when observed in more than one plane as this provides evidence that the pathology is not noise but rather a three-dimensional pattern [13], [14]. Obtaining 3D volume data can be prohibitively expensive and impractical in resource-constrained scenarios, however. An alternative is scanning the brain through multiple 2D planes. Then, the key challenge becomes how to effectively capture inter-plane correlations to represent 3D features.

Existing strategies for multi-view medical image analysis are primarily categorized into two types: image-level fusion and region-level fusion. Image-level fusion techniques [15]– [18]extract and fuse features from each view, without incorporating any local correlation. These methods work effectively when ample data is available, but their performance may decline in scenarios where data is limited. This is because the true correlation of features is only a small portion compared to the entirety of the image features. To mitigate this problem, another group of methods introduce prior knowledge into the system by identifying the candidate features or regions [19]– [22]. Customarily, pre-designed candidate region of interest detection and matching algorithms are employed to capture the region-level correspondence of features. After matching features, fusion takes place. However, the detection and matching of candidate regions may be daunting for many tasks, making the pixel-/region-wise correspondence infeasible. Our pSBI detection in the cUS dataset is one such example, considering the diversity and variety of abnormal imaging patterns.

Despite the challenges mentioned earlier, we have identified an under-explored form of prior knowledge: the strong correlation between local regions around the intersecting line of two images, as shown in Fig. 1. This observation inspired us to develop a novel cross-view fusion network that leverages the geometric relationship between two intersecting images. When the spatial positions of the images are known, the location of this intersecting line can be approximately determined. Our approach specifically identifies, pairs, and fuses the correlated local regions in two views around the intersecting line, aiming to efficiently uncover cross-view local patterns. The comprehensive features extracted from the two branches, now enriched with cross-view information, are combined and input into the final classifier. Thus, we have named our proposal the Intersection-Guided Cross-view Local-and Image-level Fusion Network (CLIF-Net). The key contributions of our proposed approach include:

**Fig. 1:**
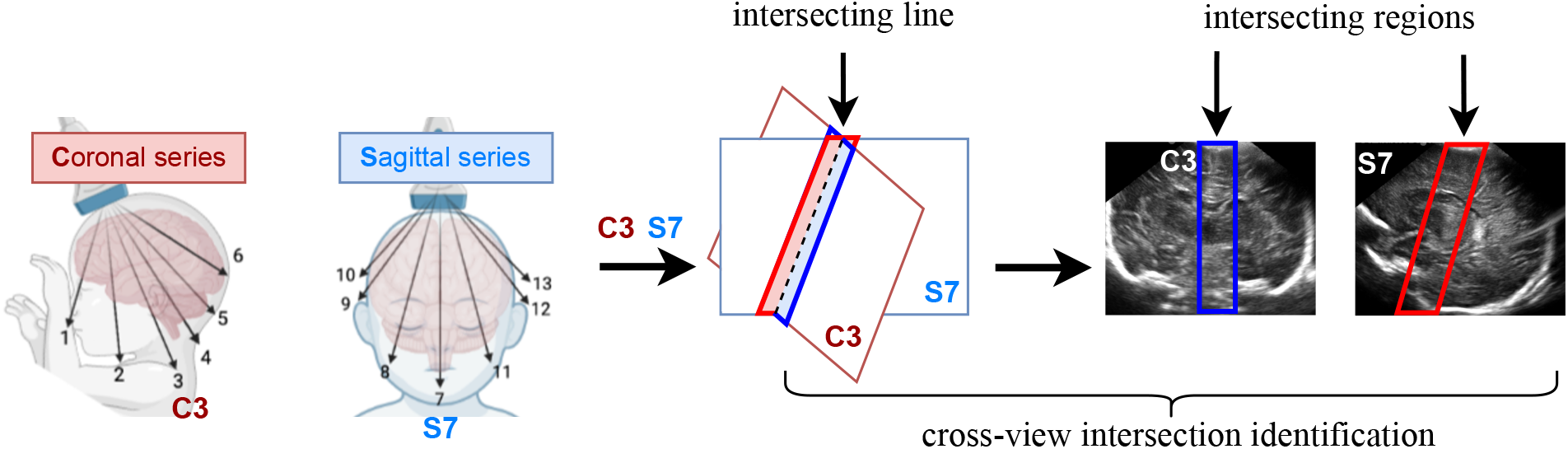
Demonstration of intersecting region from cross-view cUS images. Given the scanning position of two images, the intersecting line can be located. The intersecting region, i.e the region surrounding the line, may exhibit significant correlation between two planes, which could be utilized to form a local 3D representation.

1. **Geometric Relationship Modeling Between Multi-View Image Slices:** We introduce a new method for fusing cross-view local features by exploiting the underlying geometric relationship between intersecting images, which significantly enhances the efficacy of discovering discriminative and generalizable patterns compared to traditional attention mechanisms.
2. **An Intersection-Guided Cross-view Local-and Image-level Fusion Network (CLIF-Net)** is proposed, embodying a dual-branch architecture that utilizes the proposed intersection-guided cross-view local fusion mechanism. Coronal and saggital view images serve as the input for these branches and our work is the first to mine their cross-view geometric structure towards enabling a local feature representation that enhances classification accuracy into pSBI or healthy.
3. **New cUS Dataset and Demonstration of Training Robustness:** Experimental results on a challenging cranial ultrasound (cUS) dataset (acquired from CURE children’s hospital in Uganda) demonstrate that CLIF-Net surpasses existing state-of-the-art learning methods in pSBI identification. The benefits of CLIF-Net, owing to our exploitation of problem-specific geometry, are most pronounced in limited training setups thereby leading to marked improvements in generalizability.

## II. RELATED WORK

Similar to other image classification pipelines, brain image classification involves two key steps: feature extraction and a learned classifier. Early work focused heavily on handcrafted features. Chaplot *et al*. [23] utilized wavelets as inputs for neural network self-organizing maps and support vector machines [24], aiming at classifying magnetic resonance (MR) images of the human brain. Tabrizi *et al*. [25] proposed a method to extract morphological features of cerebral ventricles from cUS images to predict the outcome of post-hemorrhagic hydrocephalus. In recent years, deep learning has become the predominant method in brain image classification for various modalities [26]–[28].

### 1) Ultrasound

Kim *et al*. [29] demonstrated that a CNN model trained on a limited dataset can detect germinal matrix hemorrhage on cUSs with high accuracy. Another study reveals that a multi-task deep learning model can both segment white matter and predict the risk of white matter injury from cUS images simultaneously [30]. Estermann *et al*. introduced a multi-modal detection/segmentation model for punctuate white matter lesions using cUS images [31]. Valanarasu *et al*. developed a 2D cUS segmentation model under limited data [32]. Martin *et al*. developed fully convolutional networks for 3D cUS ventricular segmentation. [33]. Xie *et al*. proposed a deep CNN for detecting abnormalities in the fetal brain using standard plane cUS images [34]. Another study also shows feasibility of using deep-learning algorithms to classify as normal or abnormal sonographic images of the fetal brain obtained in standard axial planes [35].

### 2) Other Modalities

Deep learning has also been leveraged for other imaging modalities. ResNet was utilized to classify brain tumors from MRI images [36]. A hybrid 2D/3D model has been proposed to classify hydrocephalus and infection from CT brain images. Additionally, cooperation between the two branches was enforced through attention learning [37].

In brain image analysis, a single-plane view of the image is often sub-optimal as abnormalities typically manifest in 3D morphology. Ideally, collecting and analyzing 3D data would be the preferred approach [37]–[41]. However, collecting high-resolution 3D imaging data is often infeasible in medical resource-constrained environments. As an alternative, a potential solution involves capturing images from multiple planes of the region of interest, providing an approximate representation of 3D features. The primary challenge then lies in the effective fusion of complementary and correlated information from multiple planes.

### 3) Image-level Fusion

Typical image-level fusion methods concatenate global features representing different views, extracted by 2D CNNs or transformers [15]–[18], [42]. To transfer information between unregistered views at the level of spatial feature maps, Tulder *et al*. [43] introduced an innovative cross-view transformer method [44]. This approach was shown to be effective when applied to multi-view mammography and chest X-ray datasets. In a study by Pan *et al*. [45], a multi-view separable pyramid network was proposed, where representations from axial, coronal, and sagittal views of PET scans were learned and fused. Jang *et al*. [46] proposed a three-dimensional medical image classifier that employed a multi-plane and multi-slice transformer network to classify Alzheimer’s disease in 3D MRI images. The methods mentioned above primarily concentrate on global feature fusion, and they lack the capacity to simultaneously analyze and incorporate both global and local information from multiple views. To address this, Chen *et al*. [47] proposed a model for fusion of global and local features from ipsilateral views in mammograms images [48].

### 4) Region-level Fusion

Image-level fusion proves beneficial when abundant data is available, as for many tasks only a minor portion of the total features are critical. As a result, some methods employ prior knowledge, such as candidate detection algorithms, to limit the fusion region.

Yan *et al*. [49] introduced a multi-task learning model for dual-view mass matching and mass classification on mammograms, demonstrating that mass matching enhances the overall detection performance. Similarly, Ma *et al*. [50] utilized Faster R-CNN [51] to identify candidate regions from two views. Attention between candidate regions is then estimated and incorporated based on geometric and semantic similarities. Setio *et al*. [52] proposes to use pre-designed candidate region detectors to identify nodule candidates, where 2D patches from multiple planes are extracted and fused. Other methods also utilize cross-view candidate region detection to select and combine features from multiple views [19]–[22].

Although the aforementioned methods have demonstrated success in certain domains, they tend to lose their effectiveness when dealing with problems involving limited data, or when no candidate region detection algorithm is applicable. pSBI detection in cUS images is one such challenge. To the best of our knowledge, no existing methods have attempted to identify the intersecting line and its surrounding region and leverage the complementary semantic features from two cross-view images for pSBI detection. This inspired us to propose a model that harnesses the power of feature fusion from local region surrounding intersecting lines in cross-view images.

## III. METHOD: CLIF-NET

CLIF-Net is a dual-branch architecture that takes as input a pair of images and outputs a prediction score. It extracts the features from two views, discovers and encodes the correlation of local features in the intersecting region, and fuses the features to a global feature representation for prediction. The overall architecture is shown in Fig. 2.

**Fig. 2:**
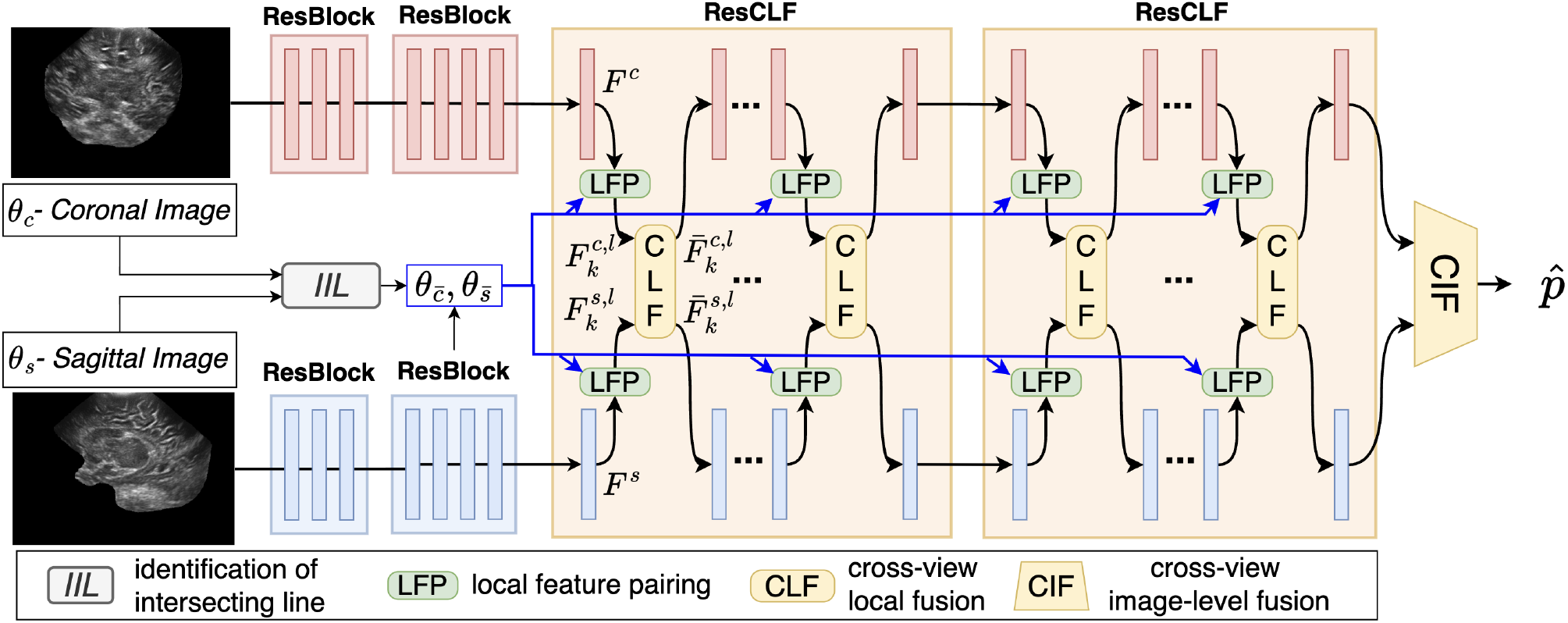
The architecture of CLIF-Net. Coronal and sagittal images serve as the input for two separate branches. With the guidance of the intersecting region, local features from the two branches interact through Cross-view Local Fusion (CLF) module. The final image-level features from both branches are fused via CIF block.

### A. Dual-branch Architecture

Each branch consists of four residual blocks (ResBlocks) [53], which sequentially reduces the dimension of the feature maps and enriches the semantic information. The first two Res-Blocks operate independently between two branches, whereas the third and fourth ResBlock enable local features interaction between branches through a Residual Cross-view Local Fusion Block (ResCLF). Differing from the conventional cross attention modules, the cross attention mechanism in proposed ResCLF focuses on the region near intersecting line – a region that exhibits a strong correlation between the two views. The specifics of the innovative block are detailed in Sec. III-B.

After the enhanced dual feature extraction process, the image-level feature tensors {**F**^*c*^, **F**^*s*^}generated by the two branches are mapped to the probability scores through the Cross-view Image-level Fusion (CIF) block, which consists of a global average pooling followed by a concatenating operation resulting in a combined image-level representation vector, two fully-connected (FC) layers with a ReLU activation function followed by a softmax output function that yields the probability scores.

The whole dual-branch network is optimized in an end-to-end manner by minimizing the binary cross entropy between 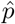 and ground-truth label *y*:

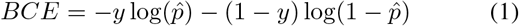

### B) Residual Cross-view Local Fusion (ResCLF) Block

Two branches’ communication is enabled by the ResCLF block, which fuses local information from two views around the intersecting line. It involves three steps: first, identifying the intersecting line; second, pairing the local features along the intersecting line; and third, fusing the pairs via a crossattention module.

#### 1) Identification of Intersecting Line

Given a pair of intersecting planes, identifying the position of intersecting line within each plane requires estimating two planes’ relative positions. All the coronal and sagittal planes in neonatal ultrasounds are actually rotated planes to the principal coronal plane and principal sagittal plane, as depicted in Fig. 3, because each ultrasound image is captured through the infant’s anterior fontanel – a bone-free area at the top of the skull. Hence, the position of each plane is described by its rotation angle relative to its principal coronal or sagittal plane, denoted as *θ*_*c*_ and *θ*_*s*_, which are hyper-parameters that can be estimated and adjusted according to specific scanning protocols. Once set, the relative position between images can be determined.

**Fig. 3:**
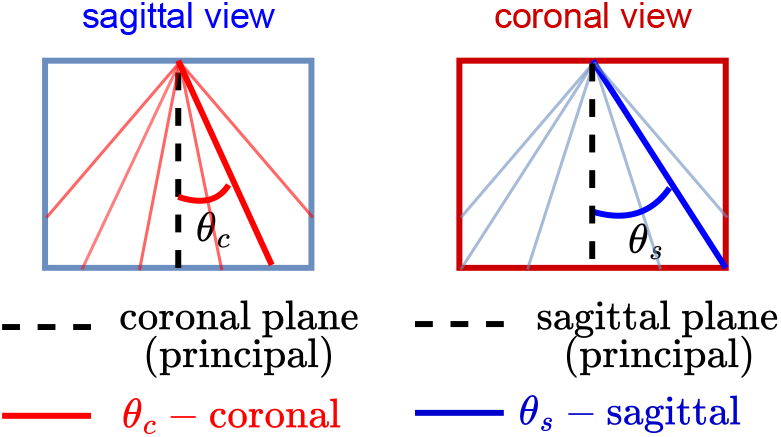
Left: coronal images (red lines) seen from sagittal view. Right: sagittal images (blue lines) seen from coronal view.

Fig. 4a demonstrates the region around the intersecting line between a *θ*_*s*_−sagittal plane (blue plane) and a *θ*_*c*_−coronal plane (red plane). Line *OB* is the intersecting line. *OA* and *OC* are the mid-lines in the *θ*_*c*_−coronal plane and *θ*_*s*_ −sagittal plane.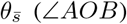 and 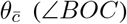 specify *OB*’s position within the *θ*_*s*_ −sagittal plane and the *θ*_*c*_ −coronal plane, respectively. From basics of geometry:

**Fig. 4:**
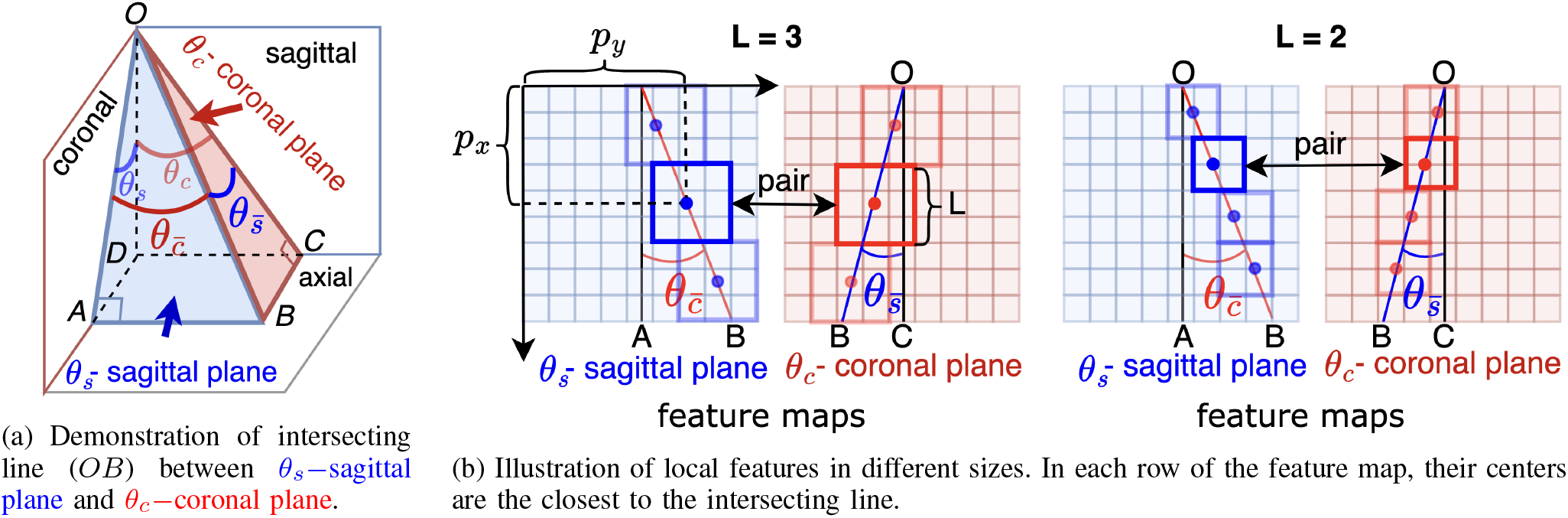
Illustration of identifying intersecting line and pairing local features.

**Fig. 5:**
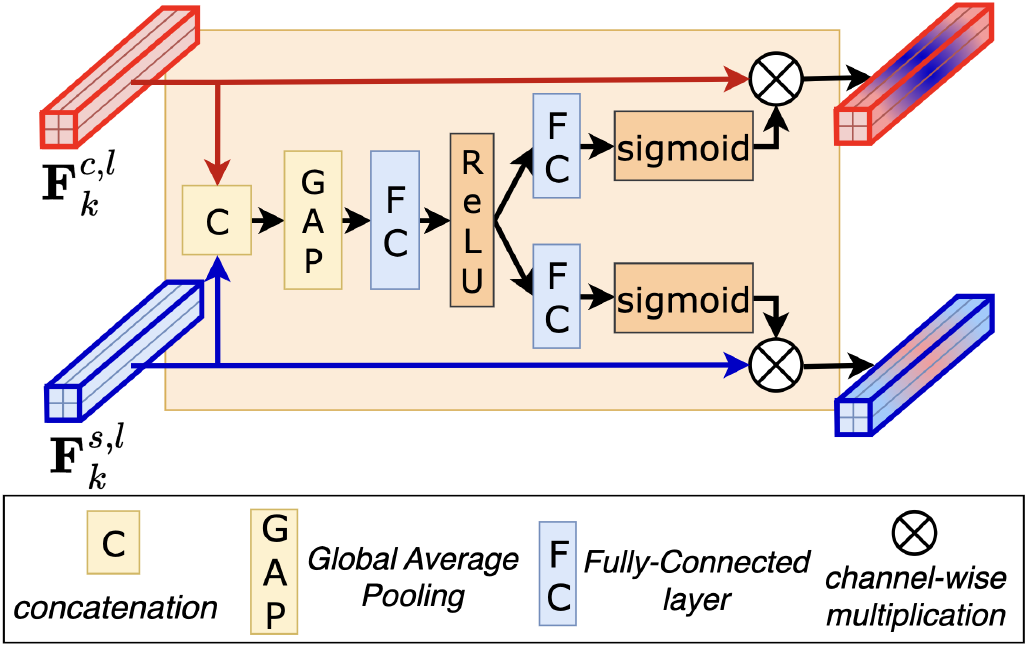
Cross-view Local Fusion (CLF) Module. The module concatenates features from coronal and sagittal planes and employs a FC layer to extract the correlated features and two separate FC layers to inject the learned correlation into the original features.

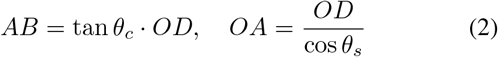

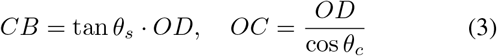

Since *BA* ⊥ coronal plane, *BC* ⊥ sagittal plane, we have:

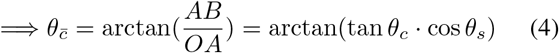

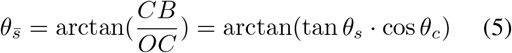

#### 2) Local features pairing

To discover cross-view local correlations, local features (LF) are extracted from the feature maps {**F**^*c*^, **F**^*s*^} through bounding boxes of shape *L*×*L*. Hence, 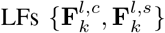 can be denoted as:

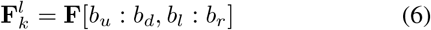

where {*b*_*l*_, *b*_*r*_, *b*_*u*_, *b*_*d*_} are boundaries of the box. To ensure local features from the same region to be paired and to be processed, the bounding boxes should follow three conditions: 1) the boxes should not overlap; 2) the boxes’ centers should locate at or approximate to the intersecting line; 3) boxes in two views have the same vertical positions.

To meet the conditions, a sequence of points following the 7 are designated as target center points of the bounding boxes. Bounding boxes’ centers should be the exact or the nearest to the target center points.

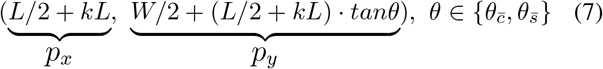

where *H, W* are the height and width of the feature maps, *k* ∈ {0, 1,…, min(⌊*H/L*⌋, *W/* ⌊*L*·*tanθ*⌋) }. The constraint on *k*’s range guarantees bounding boxes inside the feature maps.

The target center points are located at the intersecting line, which is illustrated in Fig. 4b. Therefore, bounding box’s boundary {*b*_*l*_, *b*_*r*_, *b*_*u*_, *b*_*d*_} can be derived based on those target center points in 7.

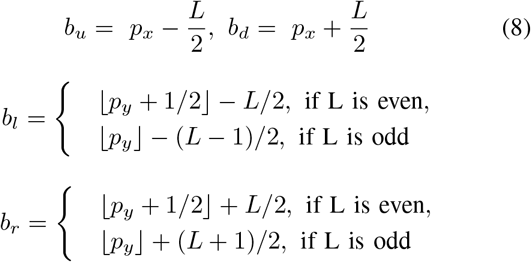

⌊·⌋ is the floor function. Following the procedures above,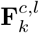 and 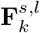 can form a pair, since they are both proxy to the intersecting line and in the similar depth (vertical distance to point *O*). The process is illustrated in Fig. 4b.

#### 3) Cross-view local fusion module

: Paired local feature tensors 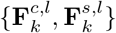 are supplied to the CLF module, which adapts the channel attention mechanism [54] to the cross-view scenario. Initially, two tensors are concatenated along the channels and then globally average pooled. This process facilitates more representative features. Then, the feature vector containing two views’ information is processed by a fully-connected (FC) layer *f*_1_(·) and a ReLU function to capture correlation between views, as shown in (9):

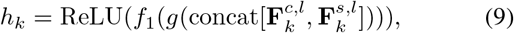

where *f*_1_(·) is FC layer, *g*(·) is global average pooling, *i* means the *i*^*th*^ residual block and *k* means the *k*^*th*^ pair. Next, the output of the FC layer is processed through two distinct FC layers, each designed to further comprehend the complex cross-view correlations and convert these correlation patterns into weights for the channels of the two local feature tensors. A sigmoid function is applied after these FC layers to constrain the values between 0 and 1, thereby enabling them to serve as weights. This operation ensures that the transformed correlations contribute to the local feature tensors in a weighted manner, which is shown in (10) and (11).

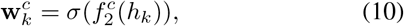

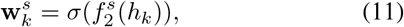

Where 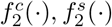 are FC layers for coronal plane and sagittal plane, respectively, and *σ*(·) is the sigmoid function.

Lastly, the weight vectors 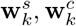 are multiplied to the local feature tensors 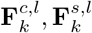 resulting in 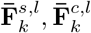, which replace the corresponding local feature tensors in **F**^*c*^, **F**^*s*^. The process is shown as follows:

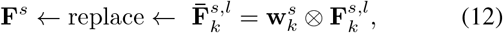

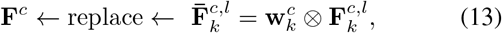

By processing the pairs of local feature tensors 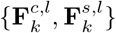 correlation of features from two views have impacts on the attention of the features. For instance, if co-existence of activation of some features from the coronal plane and some features from the sagittal plane contribute to minimize the loss, the layers in CLF module would put more weight on the corresponding features. On the contrary, if an activation of a feature contradicts several other features, less weight can be placed on that channel, which equivalently benefits selecting the most representative features for the task.

## IV. EXPERIMENTAL RESULTS

### A. Datasets

#### 1) Inclusion Criteria

We collected scans from newborn infants born at or near their expected date (term age) at the Mbale Regional Referral Hospital in Uganda. These infants were part of a large study assessing the role of cUS in aiding a diagnosis of pSBI in a low-income setting without resources for standard blood tests. The infants older than 28 days are excluded from the dataset, leading to 258 patients with clinical features suggestive of pSBI and 44 control patients. The definition of pSBI was the presence of one of the following three combinations of symptoms and signs: a) Fever (*>*37.5°C), lethargy and poor feeding; b) Hypothermia (*<*35.5°C), lethargy and poor feeding; c) Full fontanelle and/or seizures, fever and poor feeding [5].

#### 2) Ultrasound Imaging Procedures

For each patient, images are recorded using a Sonosite M-Turbo ultrasound machine and a Cx11 probe, which are performed by one of five trained clinicians. Due to hyper-echogenecity from bone, the first coronal image and left-most and right-most sagittal images are not used for the model development. For coronal view, the examination begins anteriorly and the probe is angled posteriorly. The first image demonstrates the corpus callosum, frontal horns of the lateral ventricles and temporal lobes. The second image captures the lateral ventricles, the septum pellucidum and the corpus callosum. The third image demonstrates the parietal lobes, basal ganglia and thalami, the lateral ventricles and corpus callosum. The fourth image captures the trigone of the lateral ventricles and the choroid plexus. The fifth image displays the parieto-occipital lobes. **The third plane is set as the principal coronal plane**.

For sagittal view, the examination begins in the mid-line, capturing the corpus callosum and cerebellar vermis as an echogenic image in the posterior fossa. The second and the third images are undertaken bilaterally through the right and left lateral ventricles. The fourth and fifth images visualize the white matter adjacent to the lateral ventricles out to the Sylvian fissure. **The mid-line plane is set as the principal sagittal plane**. Details of the cUS scanning protocol can be found in [5].

#### 3) Plane Angle Estimation

Angles associated with the coronal planes and sagittal planes {*θ*_*c*_, *θ*_*s*_ } can be estimated and adjusted according to certain ultrasound scanning procedures. For our dataset, {*θ*_*c*_}’s are set to be {−30^*°*^, −15^*°*^, 0^*°*^, 15^*°*^, 30^*°*^}.{*θ*_*s*_}’s are set to be {−30^*°*^, −15^*°*^, 0^*°*^, 15^*°*^, 30^*°*^}.

### B. Experimental Setup

#### 1) Training-Validation Configuration

5-fold cross-validation is adopted for comprehensive evaluation. In each fold, 206 clinical pSBI patients and 36 control patients are used for training. 52 pSBI patients and 8 control patients are evaluated.

#### 2) Skull Stripping

Skull stripping is a key pre-processing step. Irrelevant signals such as hands holding the infant’s head may provide artifact. Inappropriately angled probe technique may also lead to increased artifact from bone adjacent to the fontanelle which can hinder higher resolution imaging of the underlying brain structures. Existing skull stripping methods in cUS [34], [55], [56] primarily focus on axial views rather than coronal and sagittal views because the brains are not fully bounded by the skull in these views. Therefore, we developed a U-Net-based [57] skull stripping model specifically for coronal and sagittal neonatal cUS images, which ensures the exclusion of any extraneous information outside the brain.

First, dataset for skull stripping is created. Binary brain masks of 528 coronal images and 616 sagittal images from 44 pSBI patients (randomly selected) and 44 controls were manually labeled using Matlab imaging processing annotation (MRP, TH). We split our labeled dataset into two sets. Each one contains 22 pSBI patients and 22 normal controls. Two models are trained separately on each set and tested on the other. Training loss is dice loss, which is defined in 14.

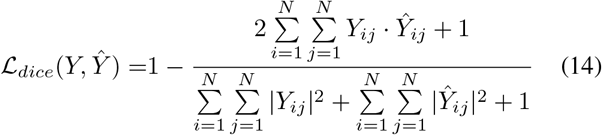

The learning rate of the network was initially set to 0.001 and was decreased by a factor of 5 every 20 epochs. The Adam optimizer was adopted for optimization [58]. The network was trained for a total of 100 epochs, with a batch size of 16. During training, data augmentation techniques such as rotation, flipping, and scaling were applied. Cross-evaluated on the two labeled datasets, skull stripping model can achieve 98% dice score.

An example of skull stripping output and ground-truth is shown in Fig. 6. After training, each patient’s images in the dataset are skull stripped using one of two models (Model A or Model B). If a patient is included in the training dataset of Model A, Model B is used; if a patient is included in the training dataset of Model B, Model A is used. If a patient is not included in either model’s training dataset, one of the models is randomly assigned.

**Fig. 6:**
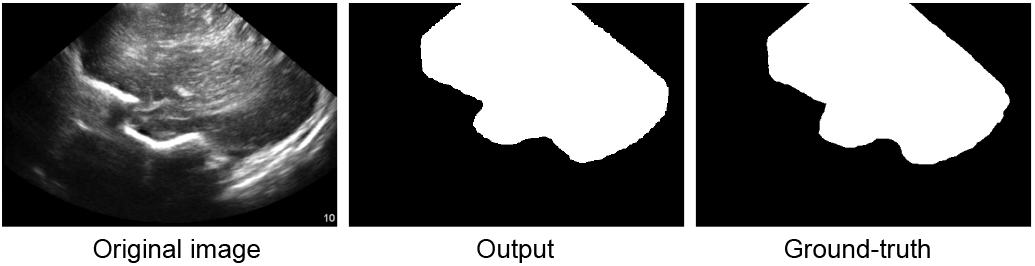
Results of skull stripping network. Left: original images; middle: output masks; right: ground-truth masks.

#### 3) CLIF-Net

The overall architecture and loss function of CLIF-Net are described in Sec. III and detailed in Tab. I. The local feature width *L* is set to 6 in the ResCLF1 and 3 in the ResCLF2, which are determined via cross-validation [59]. The learning rate of the network was initially set to 0.0001. The adaptive learning rate strategy is adopted here, where the learning rate was decreased by a factor of 5 when the validation loss has not decreased for 10 epochs. The optimizer used here was the Adam optimizer. The network was trained for a total of 200 epochs, with a batch size of 16. Augmentation in Sec. IV-B.2 is also applied during training. In addition, the angles *θ*_*c*_, *θ*_*s*_ are also augmented by adding a noise to simulate the angle estimation noise in real-world clinical application. **Inference**: top 5 most confident scores from outputs of 25 pairs of one patient are averaged as the final prediction score.

#### 4) Evaluation

In the evaluation phase, pSBI patients are defined as positive cases. Non-pSBI patients are defined as negative cases. For the quantitative assessment of CLIF-Net and the comparison with other methods, we use sensitivity (SEN), specificity (SPE), and F1 score (F1).

### C. Ablation Study

In this section, we perform ablation studies for our proposed method to assess the impact of the components of CLIF-Net.

#### 1) Impact of Skull Stripping

First, we investigated the impact of skull stripping to the classification capability. As we can see from Table II, the specificity significantly drops without skull stripping even though the sensitivity increases slightly. This is due to the overfitting of similar signals outside skull of control patients that lead to predicting pSBI. Skull stripping eliminates potential confounding features outside of the skull. In addition, removing the irrelevant signals is beneficial to cross-view local fusion by filtering out distracting information.

**TABLE I:**
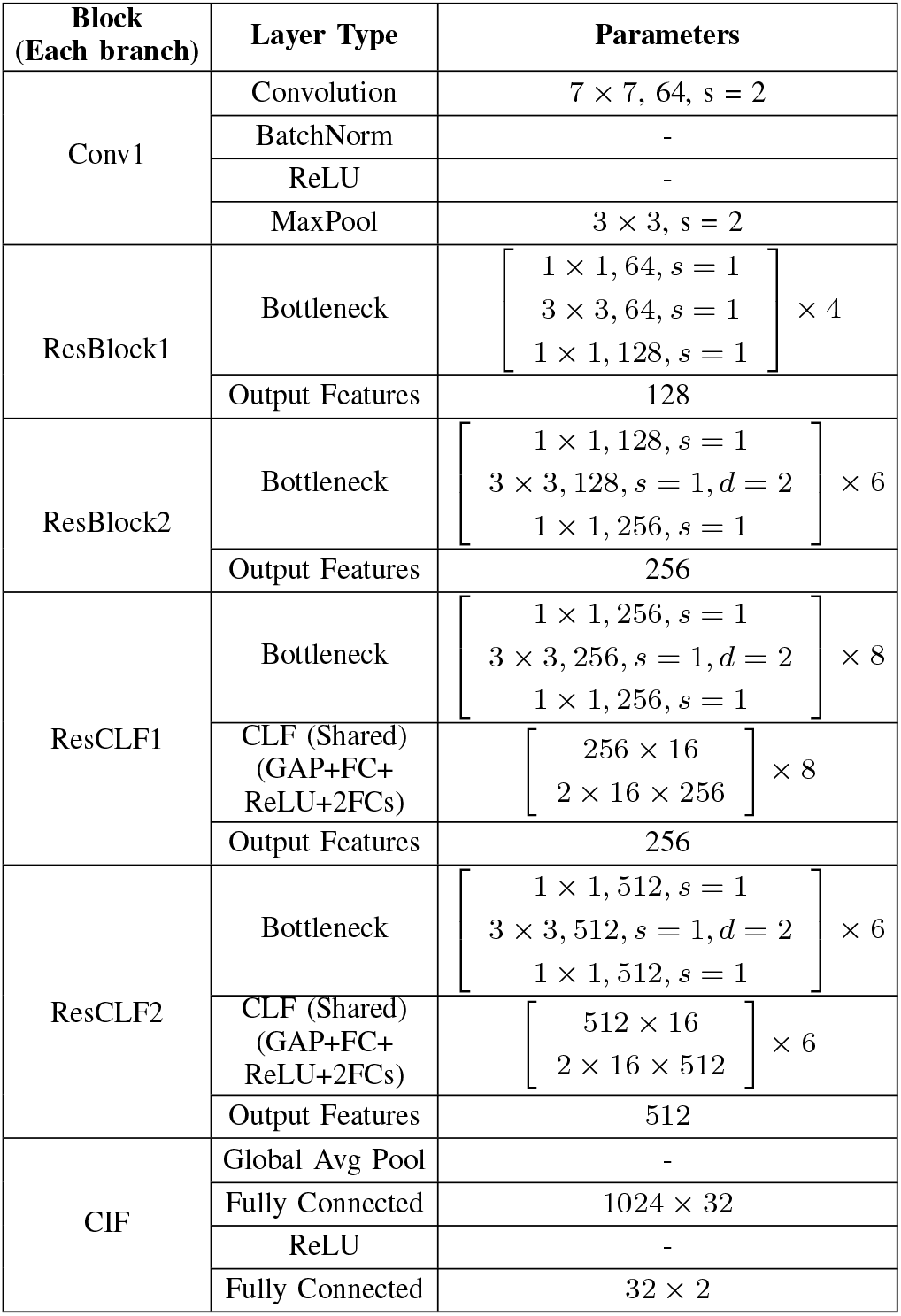
Block-wise parameters (kernel, filters, stride, dilation) for CLIF-Net. In the column for parameters, kernel size, number of filters and stride are reported.

**TABLE II:** Ablation study of skull stripping’s impact to the classification results.

|  | Sensitivity | Specificity |
| --- | --- | --- |
| without skull stripping | 0.923 | 0.612 |
| with skull stripping | 0.912 | 0.864 |

GradCAMs [60] of a control patient is shown in Fig. 7. It can be seen that there is large activation outside the brain region when the model is not trained with skull-stripped images, causing a reduction in specificity.

**Fig. 7:**
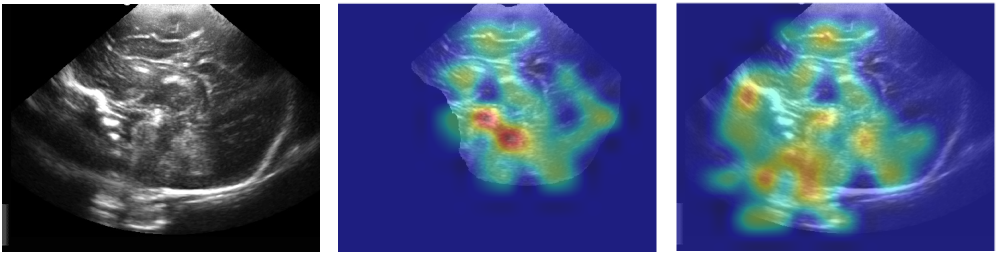
Left: original US image. Middle: CAM from CLIF-Net trained on skull-stripped data. Right: CAM from CLIF-Net trained on original data.

#### 2) Effectiveness of CLF and CIF

We investigate the effectiveness of CLF and CIF by following experiments: A) neither CLF nor CIF is incorporated, resulting in a completely single-view method with no communication between views. Majority voting is adopted to obtain final prediction. B) The CIF block is incorporated, allowing for image-level fusion. Two branches have no interaction. C) Both CLF and CIF are incorporated, enabling both local and image-level fusion. However, the inputs in CLF are randomly paired. D) Both CLF and CIF are incorporated, enabling both local and image-level fusion.

We then analyze the results to understand the impact of CLF and CIF on the model’s performance. The sensitivity, specificity, and F1 score can be found in Table III. Additionally, we conducted the Mann-Whitney U test [61] between the F1 scores of experiments A, B, C, and D.

**TABLE III:** Ablation study of CLF and CIF blocks. Comparison of B-D and C-D indicates that learning local patterns in intersecting region benefits the overall cross-view model.

| Exp. | CLF | CIF | SEN | SPE | F1 | *p*-value (F1) |
| --- | --- | --- | --- | --- | --- | --- |
| A | × | × | 0.864 | 0.818 | 0.910 | 0.00044 |
| B | × | ✓ | 0.887 | 0.835 | 0.926 | 0.031 |
| C | ✓ (Random) | ✓ | 0.872 | 0.825 | 0.921 | 0.017 |
| D | ✓ | ✓ | **0.912** | **0.864** | **0.942** | - |

The findings indicate that incorporating the CIF block enhances performance. Notably, significant improvement over experiment B is observed when CLF is incorporated, showcasing the effectiveness of capturing cross-view features from the local region around intersecting line.

To further illustrate the impact of CLF, we present grad-CAMs [60] from CLIF-Net with and without CLF blocks in Fig.8. These images are derived from two patients with pSBI. Without the CLF module, attention is uniformly distributed over the brain region, not focusing on abnormalities, resulting in lower probability scores for both cases (0.672 and 0.598). Conversely, with the CLF block enabled, the pSBI patterns are detected through local corrections in two views. In the left pair, notable features include a widened hyperechoic interhemispheric fissure and hyperechoic sulci. In the right pair, a hyperechoic region adjacent to the choroid plexus, possibly indicative of hemorrhage or inflammation, is prominently highlighted in both views [13], [14].

**Fig. 8:**
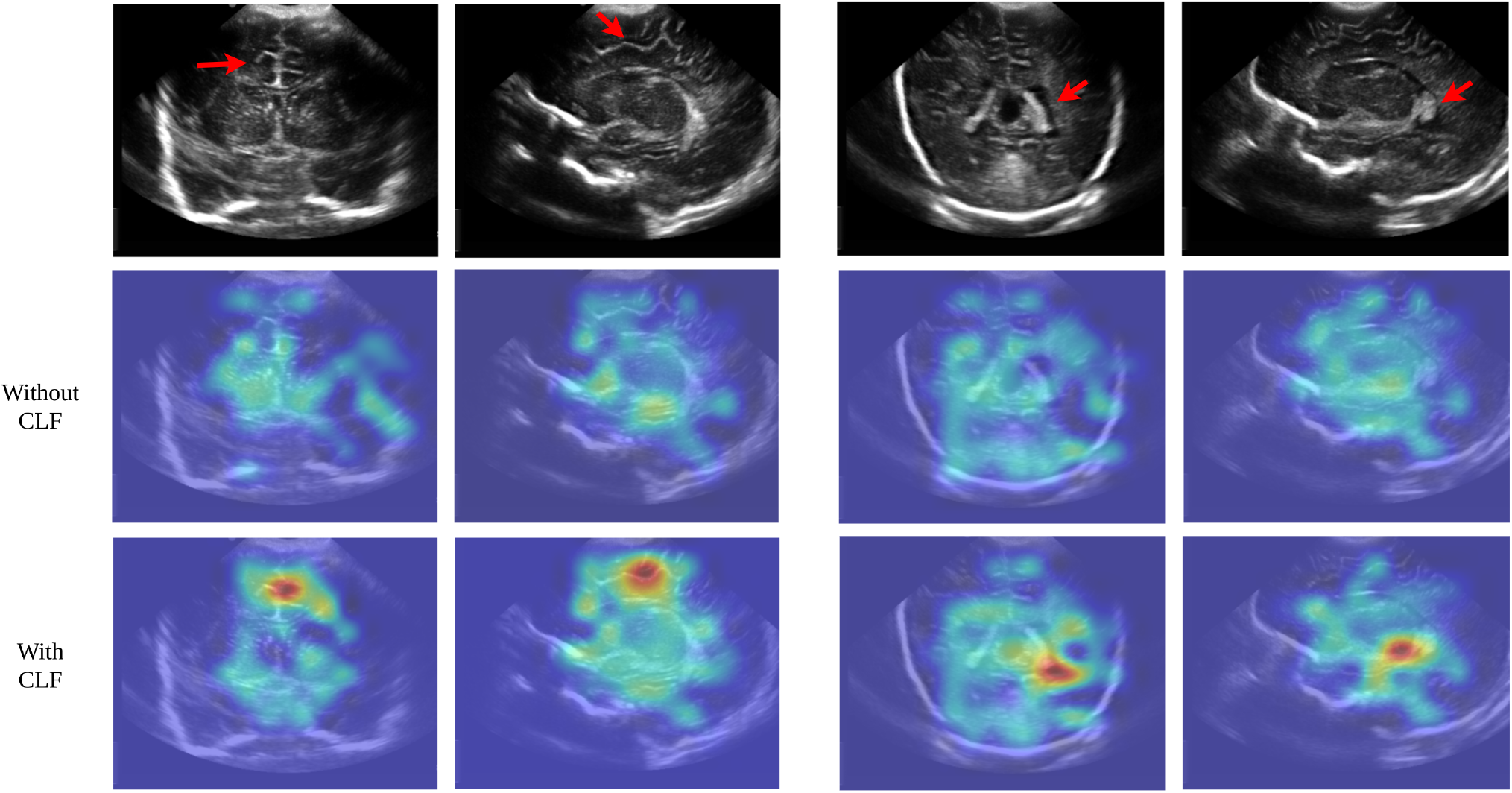
Image pairs from two pSBI patients with GradCAMs from CLIF-Net with and without CLF blocks are shown. Images in the second row are CAMs without CLF. Images in the third row are CAMs with CLF. Through cross-view learning in CLF, hyperechoic interhemispheric fissure with hyperechoic sulci are focused with more attention. Probability score for pSBI is boosted from 0.672 to 0.912 (left) and from 0.598 to 0.867 (right).

#### 3) Impacts of the CLF Module Positions and Local Feature Size L

The CLF module can be integrated into any Residual Blocks to create a ResCLF block. However, it is crucial to strategically position the CLF module within appropriate blocks, as the receptive field (RF) of each block (i.e. the region in the input image to be seen by a feature pixel) progressively expands. Given the input size to be 382 × 512, each block of our dual-branch architecture receives information from different aspects:

- 1^*st*^ block: the RF grows from 21 to 37. This block primarily processes local textural and edge information from the image.
- 2^*nd*^ block: the RF grows from 37 to 85. It can capture larger features such as small cysts, lesions, or localized inflammation.
- 3^*rd*^ block: the RF grows from 85 to 149. This block captures substantial regions of the ultrasound image, allowing it to interpret larger anatomical structures within a broader context. It can recognize significant abnormalities that span larger areas, such as widespread edema or larger infected regions.
- 4^*th*^ block: the RF grows from 149 to 197. The largest receptive field in this block allows the network to analyze substantial portions of US images. This capability is critical for making overarching assessments of the cranial space, correlating various signs of infection or other abnormalities across the entire image.

Therefore, we investigated CLIF-Net’s performance when the CLF module is integrated within different residual blocks of the dual-branch architecture. Additionally, for each position, we conducted experiments with different local feature sizes, as varying the size may impact the representation capacity for local information.

From the results presented in Table IV, it is evident that placing the CLF module in block 1 and block 2 is suboptimal, regardless of the local feature size. Integration into block 3 or block 4 yields superior performance compared to block 1 and block 2. This observation suggests that fusing mid-to high-level features is more effective for our model. Additionally, this could be attributed to the increased robustness of local feature extraction and pairing in deeper layers due to the larger receptive field.

**TABLE IV:** The average F1 score under different positions of ResCLF and size *L* is reported here.

| Block to integrate CLF | $L$ / F1 | $L$ / F1 | $L$ / F1 |
| --- | --- | --- | --- |
| **Block 1** | 8×8 / 0.914 | 24×24 / 0.912 | 40×40 / 0.912 |
| **Block 2** | 4×4 / 0.923 | 12×12 / 0.925 | 20×20 / 0.920 |
| **Block 3** | 2×2 / 0.931 | **6×6 / 0.938** | 10×10 / 0.927 |
| **Block 4** | 1×1 / 0.931 | **3×3 / 0.936** | 5×5 / 0.930 |

Furthermore, when setting the local feature size *L* = 6 in block 3 and *L* = 3 in block 4, we observe improved performance compared to smaller or larger local feature sizes. We conjecture that extracting features that are too small may not capture enough information from the local region, thereby increasing the risk of over-fitting non-generalizable correlation patterns in the training data. Additionally, setting *L* too small may amplify the impact of the plane angle estimation error. Conversely, if the local feature size is too large, it may include more uncorrelated information than the CLF can effectively model. Therefore, choosing an intermediate size not only captures sufficient information and enhance the robustness to angle noise but also avoids incorporating redundant information.

#### 4) Robustness to Plane Angle Estimation Noise

Further-more, we investigated CLIF-Net’s robustness to the plane angle estimation noise, which is unavoidable in the scanning process. This study is crucial for evaluating its impact to the cross-view local feature fusion. To mimic the estimation errors in practical scenarios, a series of experiments are conducted in this study, where angle deviations (noise) *ϵ*_*θ*_’s of various levels are introduced to the testing data whereas the training data remain the same. Model’s performance is examined to illustrate its robustness under these scenarios.

In each experiment, training data are the same i.e. model remains the same. For the testing data, coronal images of each patient are associated with a deviated angle *θ*_*c*_ + *ϵ*_*θ*_ or *θ*_*c*_ − *ϵ*_*θ*_ with probability of 50%. Similarly, each sagittal image is associated with a deviated angle *θ*_*s*_ + *ϵ*_*θ*_ or *θ*_*s*_ − *ϵ*_*θ*_ with probability of 50%.

From the Table V, it can be seen that CLIF-Net is insensitive to angle noise at the level of 5^*°*^. Even when noise is 10^*°*^, the impact is still small. The degradation only becomes non-trivial when angle noise reaches 15^*°*^ or greater. When noise reaches 25^*°*^ which is almost equivalent to full mismatch between two views, the performance degrades to the same level as CLIF-Net without CLF blocks. Overall, results show that CLIF-Net is robust to small and medium angle estimation noise and still yields comparable results to image-level fusion method when noise is large. The robustness is attributed to the large receptive field of the local features and proper angle augmentation in the training process.

**TABLE V:** Angle noise is added to the testing data to test CLIF-Net’s robustness to plane angle estimation errors.

| Angle noise ($\epsilon_\theta$) | 5° | 10° | 15° | 20° | 25° |
| --- | --- | --- | --- | --- | --- |
| sensitivity | 0.912 | 0.908 | 0.898 | 0.886 | 0.880 |
| specificity | 0.864 | 0.864 | 0.853 | 0.842 | 0.837 |
| F1 | 0.942 | 0.939 | 0.933 | 0.928 | 0.922 |

### D. Robustness to Training Data Selection

Demonstrating the robustness and generalizability of our model is essential and challenging. Whereas many models perform well with abundant training data, their true effectiveness is often revealed under low-data conditions, where differences in feature extraction capability and generalizability become apparent. This is especially relevant in medical imaging, where limited data is a frequent challenge. To address this, we assessed the performance of CLIF-Net with reduced training data, benchmarking it against state-of-the-art (SOTA) methods [42], [45]. This evaluation is critical as it underscores the model’s capacity to identify the most salient and indicative features in data-scarce scenarios, thereby enhancing its ability to generalize to new, unseen data. As illustrated in Fig. 9, both the F1 score and AUC-PR of CLIF-Net demonstrate a more modest reduction compared to SOTA methods when restricted to only 70% and 40% of the training data. This emphasizes the generalization ability by exploiting the cross-view data structure i.e. integrating intersection information into the cross-view fusion process.

**Fig. 9:**
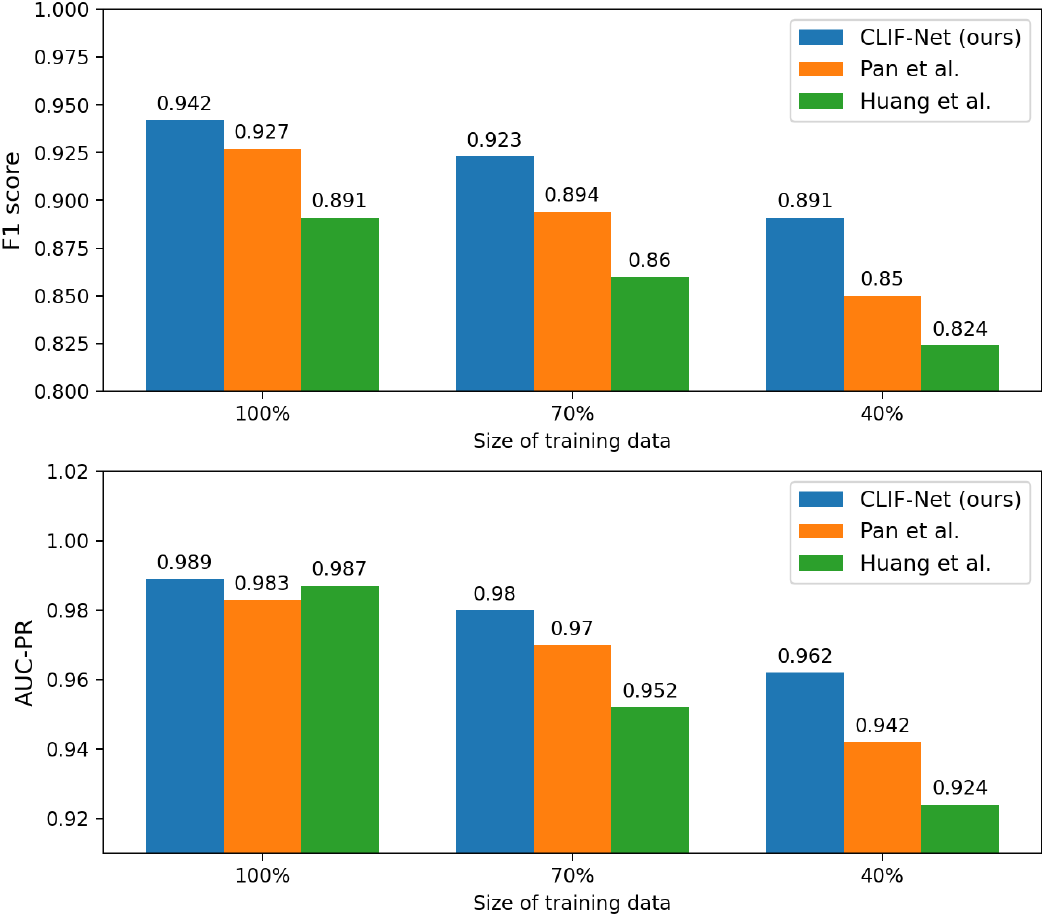
Comparison with SOTAs under the scenario of extremely low training data.

### E. Different Strategies for Outputs Ensemble

We experiment with three ensemble strategies for the final decision. Strategy A uses majority voting, whereas Strategy B selects the pair with the highest confidence. Strategy C averages the top 5 pairs with the highest confidence (probability score) to obtain the final score. Strategy D averages the top 10 pairs, and Strategy E averages the outputs of all pairs. As shown in Table VI, averaging the top 5 confident pairs achieves the best performance. Imaging patterns indicative of infection may appear in one or multiple local areas. Relying solely on the single pair with the highest probability score decreases specificity, suggesting a possible risk of overfitting towards the pSBI class. On the other hand, averaging all pairs reduces sensitivity, indicating that a significant number of regions may not contain indicative imaging patterns, which, when averaged, lowers the model’s detection capability. As a result, selecting the top 5 pairs with the highest confidence is a more sensible approach to balance the model’s detection ability and robustness against overfitting.

**TABLE VI:**
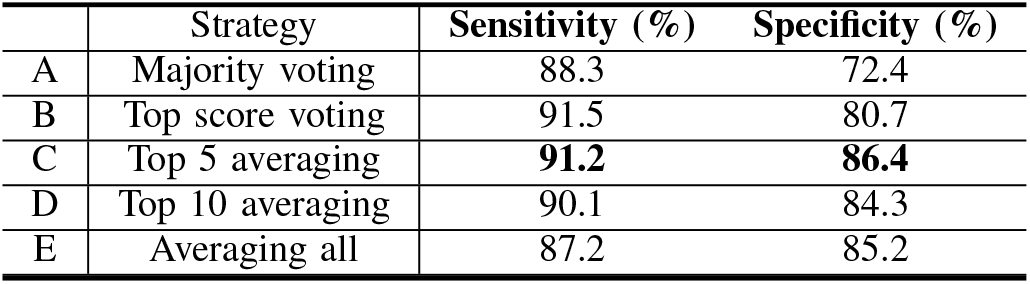
Experiments of different ensemble strategies.

### F. Robustness to Potential Domain Shift

In real-world clinical applications, deep learning-based models can be sensitive to domain shifts caused by different equipment or inconsistent imaging acquisition procedures. To simulate one type of potential shift, we applied gamma correction with varying contrast levels to the validation sets as shown in Fig. 10, while training the model without incorporating contrast adjustments. The results are presented in Table VII.

**Fig. 10:**
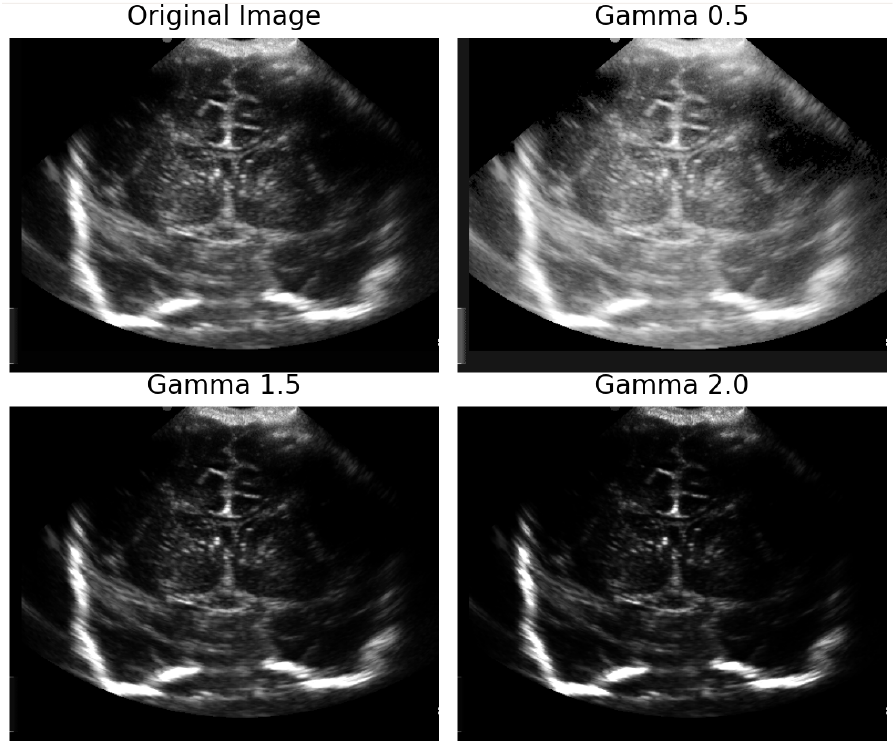
US images applied with gamma correction at different levels (*γ* = 0.5, 1.5, 2).

**TABLE VII:** Experiments of simulating domain shift via gamma correction.

| Gamma ($\gamma$) | Sensitivity (%) | Specificity (%) |
| --- | --- | --- |
| 0.5 | 91.2 | 85.6 |
| 1 | 91.2 | 86.4 |
| 1.5 | 89.4 | 82.8 |
| 2 | 86.1 | 81.8 |

The results indicate that the model’s performance degrades reasonably when gamma correction is applied, whether the simulated images are darker or brighter. This suggests that the model’s feature extraction focuses on local anatomical structures rather than intensity distribution, demonstrating its robustness to potential domain shifts in imaging acquisition. In future studies, further experiments such as collecting more clinical data or synthesizing data via generative models can be conducted to further validate model’s robustness to potential domain shift.

### G. Comparison with State-of-the-Art (SOTA)

In this section, we compare CLIF-Net against several SOTA methods for image classification, which are as follows: 1) Kim *et al*. [29]: A CNN-based single-view model. 2) Van *et al*. [56]: A dual-branch architecture with a cross-view transformer for image-level fusion. 3) Huang *et al*. [42]: Utilizes a swintransformer to extract features and fuse them via a weighting network. 4) Chen *et al*. [47]: Utilizes a self-attention module to model the correlation between two views. 5) Pan *et al*. [45]: A CNN-based cross-view model. Note that all the SOTA methods we compare against are image-level fusion methods, as there are no candidate detection algorithms that we are aware of specifically designed for pSBI detection. Additionally, we present the classification results of a human expert for 30 pSBIs and 30 normal controls. The experiment was designed so that the expert was blind to the diagnostic information.

#### 1) Discussion of Results

Table VIII presents the average values of sensitivity, specificity, F1 score, and AUC-PR over 10 repeated experiments. Mann-Whitney U test on the F1 scores is also shown in the last column. The box plot of F1 scores is displayed in Fig. 11. From Table VIII, our approach significantly outperforms single-view methods and consistently exceeds cross-view methods in both sensitivity and specificity. This highlights CLIF-Net’s ability to capture septic features effectively without overfitting, even when training data is limited. In contrast, other methods dependent on global fusion tend to overfit without sufficient local guidance and require more data. The findings indicate that learning-based models hold significant promise in using cranial ultrasound for pSBI diagnosis, and are able to exploit subtle visual cues that may be hard to identify for even a human expert. Coupled with the activation maps displayed in Fig. 8, the proposed CLIF-Net emerges as a leading and clinically valuable method for the challenging task of pSBI detection.

**TABLE VIII:** Experimental results of SOTA methods and ours on cUS dataset. p-value of Mann-Whitney U test on F1 scores between each method and our method are shown in the last column.

| Category | Methods | Sensitivity | Specificity | F1 | AUC-PR | p-value (F1) |
| --- | --- | --- | --- | --- | --- | --- |
| 2D CNN | Kim *et al.* [29] | 0.840 | 0.819 | 0.901 | 0.984 | 0.00018 |
| Cross-view transformer | Huang *et al.* [42] | 0.822 | 0.841 | 0.891 | 0.987 | 0.00018 |
| Cross-view transformer | Van *et al.* [43] | 0.862 | 0.853 | 0.914 | 0.978 | 0.0028 |
| Cross-view CNN | Chen *et al.* [47] | 0.887 | 0.805 | 0.923 | 0.982 | 0.0103 |
| Cross-view CNN | Pan *et al.* [45] | 0.899 | 0.799 | 0.927 | 0.983 | 0.037 |
| - | CLIF-Net (Ours) | **0.912** | **0.864** | **0.942** | **0.989** | - |
| Estimated performance from human expert |  | 0.800 | 0.600 | 0.727 | - | - |

**Fig. 11:**
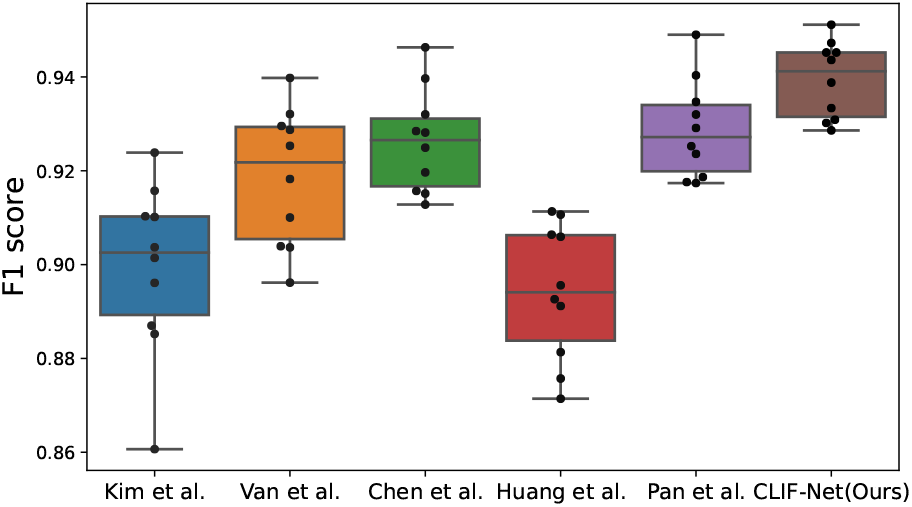
Box plot of F1 scores of SOTA methods with ours.

## V. CONCLUSION

We develop a cross-view fusion network capable of classifying multi-view cUS images. Specifically, we devise a novel method to identify the intersecting region within each individual view and develop a local fusion module to capture the correlations between the views. Experimentally, our proposed network has demonstrated its effectiveness through higher F1 scores compared to existing methods. The ability to capture and exploit information from the intersecting regions has proven to be valuable in achieving improved performance in medical image classification tasks.

In our application, we have shown that this method can discriminate neonates with pSBI from controls using multiview cUS. It is important to note that the differences seen between the brain scans of the control infants and those with pSBI likely represent a systemic pro-inflammatory state that includes increased blood-brain barrier permeability and neuroinflammation [62]. Not all of the pSBI infants in this study had clinical or imaging findings consistent with specific and singular neuroanatomical infections such as meningitis, encephalitis or ventriculitis. This posits that the generalized inflammation secondary to increased innate immune system response and cytokine storm contributes to patterns of abnormal echoic signal that are differentiable between pSBI and healthy control infants [63]–[65]. Such pro-inflammatory states are not specific to bacterial infection, but can be present in fungal, parasitic, or viral settings as well. Since most neonates suffering from possible severe bacterial infections are in settings without adequate diagnostic laboratories, our method could be used to support the development of a novel strategy for rapid optimization of diagnosis and therapy at point-of-care for this life-threatening condition. In the future, it could be worthwhile to investigate forming a detection task for precise localization of infection and inflammation and collect more data to explore the feasibility of 3D volume reconstruction for more precise analysis.

## Data Availability

All data produced in the present study are available upon reasonable request to the authors

